# Treatment-Structured Modeling of Tuberculosis Transmission with Threshold Dynamics, Stability Analysis, and Implications for Disease Control

**DOI:** 10.64898/2026.07.28.26359108

**Authors:** Jannatun Nayeem, M. A. Salek, M. H. A. Biswas, M. Humayun Kabir

## Abstract

**Background:** Tuberculosis remains a persistent infectious disease whose control is complicated by latent infection, delayed treatment, incomplete recovery, reinfection, and continuing transmission from infectious individuals. Although treatment is central to tuberculosis management, it is frequently represented only as a transition parameter in mathematical models rather than as a separate epidemiological state. In this study, treatment was therefore incorporated explicitly as an independent compartment so that its influence on transmission, recovery, disease-induced mortality, and long-term disease persistence could be evaluated.

**Methods:** A deterministic nonlinear compartmental model was formulated by dividing the total population into susceptible, exposed, actively infected, treated, and recovered classes. Reinfection of recovered individuals, progression from latent infection to active disease, movement of infectious individuals into treatment, treatment-associated recovery, natural mortality, and disease-induced mortality were included. Positivity and boundedness of the solutions were examined to establish biological validity. The basic reproduction number, *R*_0_, was derived through the next-generation matrix approach. Disease-free and endemic equilibria were determined, and their local and conditional global stability properties were investigated using Jacobian analysis, the Routh–Hurwitz criterion, center manifold theory, Lyapunov functions, and LaSalle’s invariance principle. Normalized sensitivity indices, Latin hypercube sampling, partial rank correlation coefficients, and numerical simulations were also applied.

**Results:** The disease-free equilibrium was shown to be locally asymptotically stable when *R*_0_ < 1, whereas sustained transmission and a unique endemic equilibrium were associated with *R*_0_ > 1. Under the stated reduced-model assumptions, stability of the endemic equilibrium was established. Transmission-related parameters were identified as the strongest positive contributors to disease persistence. In contrast, treatment and recovery parameters were found to reduce the reproduction number and infectious burden. Numerical simulations indicated that stronger treatment implementation and reduced transmission opportunities produced substantial reductions in active tuberculosis cases.

**Conclusion:** Treatment was shown to function as both a clinical pathway and an epidemiological control mechanism. The proposed framework may support the design of treatment-centered strategies for reducing tuberculosis prevalence and preventing long-term endemic persistence.

## 1. Introduction

Tuberculosis (TB) is an infectious disease caused by the bacterium *Mycobacterium tuberculosis*.The increasing prevalence of tuberculosis has emerged as a significant global health issue. This disease can affect various human organs. Consequently, individuals who inhale air tainted with the bacteria are at risk of contracting tuberculosis (Senedu B. Gebreegziabher, 2016). If a person is infected with TB, the bacteria may not be directly active and may not make them sick. These people have no symptoms and cannot transmit TB to others. Then we talk about latent TB. When individuals are exposed to Mycobacteria tuberculosis through sputum from infected people, it represents a latent stage, when the bacteria in the body are inactive due to the activity of the immune system. Therefore, latent individuals cannot transmit the disease to others. Usually, early TB infection occurs in the asymptomatic stage, when the infection is asymptomatic. If their immune system is weak and they do not receive proper treatment, latent tuberculosis (TB) has the potential to progress into active tuberculosis. The manifestations of active TB include persistent coughing that may involve blood for three weeks or longer, significant weight loss, extreme fatigue, elevated body temperature, and episodes of sweating during the night (Azouaoui Mbarek, 2024). The primary reason for the high number of TB cases globally is the failure of treatment due to a lack of compliancewith the prescribed treatment and a lack of knowledge among TB patients. If the patient is not treated according to the doctor’s advice then the risk of drug resistance may also increase (Holly A. Taylor, 2022). In 2022, WHO implemented the Directly Observed Treatment (DOT) program for the treatment of tuberculosis.DOT involves directly observing or monitoring tuberculosis patients while they take anti-tuberculosis medicines to confirm the correct combination and dosage of the drugs. Patients are monitored daily by healthcare workers or trained individuals who are knowledgeable about the use of medication.The DOT strategy can reduce the spread of tuberculosis more cost-effectively(Siti Aishah Abas, 2024). People with tuberculosis can continue to live healthy lives for months without any symptoms. During this time, the disease caused by the bacteria “Mycobacteria tuberculosis” (MTB) will be transmitted. The mode of transmission is usually by the patient coughing up sputum into the environment or by the bacteria. But in cases where the immune system is unable to do this, droplets containing the bacilli will be dispersed when coughing. Shows sufficient resistance, the tuberculosis bacteria become although tuberculosis is an infection that can occur in more active people and tuberculosis occurs(Mary Lilián Carabalí-Isajar, 2023). Transmission occurs through the air containing tubercle bacilli by the expulsion of sputum from pulmonary tuberculosis victims when coughing, sneezing or singing. Treatment of tuberculosis becomes difficult due to the presence of multidrug-resistant tuberculosis. Treatment takes longer and requires more expensive drugs. But for many developing countries, the maintenance is completely free.

A mathematical model is a mathematical representation that describes the behavior of a real-world problem, one of which is the spread of infectious diseases. Mathematical modelling is a field of mathematics that allows real-world problems to be represented and explained in mathematical language and thus gain a more accurate understanding of a real-world problem(Piotr Wąż 2, 2024). Mathematical modeling has demonstrated its efficacy as a valuable instrument for examining the transmission dynamics of infectious diseases.

Numerous investigations have been undertaken over the years to explore the dynamics of tuberculosis. In this section, we develop and analyze a fundamental mathematical model that addresses the transmission dynamics of both pulmonary and extrapulmonary tuberculosis, drawing upon existing research in the field (Abdulsamad Engida Sadoa, 2024). The World Health Organization classified extra pulmonary tuberculosis as infectious type I, while those with extra pulmonary tuberculosis as non-standard, latent tuberculosis stage and detection, malnutrition, HIV infection, alcohol consumption, smoking and diabetes mellitus pulmonary or extra pulmonary tuberculosis while the strategy against tuberculosis, two types of tuberculosis should be considered, while considered human infections due to endogenous reactivation represented by susceptible S bacteria, latently infected L tuberculosis(Md Abdul Kuddus, 2022). Numerous researchers have engaged in the study of mathematical modeling to gain insights into the transmission dynamics of various diseases (Tauhidul Islama, 2024), including tuberculosis. The study conducted by the authors (Md Abdul Kuddus, 2022)analyzed the spread of tuberculosis over recovery time. The impact of tuberculosis resurgence can be modeled using a mathematical approach like SEIR with a linear infection function. A SEIR model is employed to examine both the LSA (local asymptotic stability) and GSA (global asymptotic stability) of the tuberculosis transmission framework presented by the authors in(Kalyan Dasa, 2021). The authors of(Tauhidul Islama, 2024) developed their tuberculosis model by considering the impact of media perception on the population. The authors of (K.L. Holloway-Kew, 2023) discussed the super infection in tuberculosis. Recently, the authors (Chung-Min Liao, 2013) proposed a mathematical model to evaluate the ability of a new tuberculosis vaccine to control tuberculosis transmission. To control the spread of tuberculosis, some mathematical models implement it as an optimal control problem, such as the study conducted by (Cristiana J. Silva, 2013) discussed the optimal control problem of tuberculosis and multidrug-resistant tuberculosis models.

The mathematical representation of tuberculosis (TB) transmission dynamics within populations, along with the development of an epidemiological model, serves as a crucial approach to comprehending the dissemination of this disease. The structure of the paper is organized in the following manner. Section 2 offers a detailed explanation of the model formulation. In Section 3, we perform a qualitative analysis of the model, assessing the basic reproduction number and investigating the local and global stability of the disease-free equilibrium. Section 4 discusses the existence of a unique endemic equilibrium point, along with a concise examination of the local and global stability associated with this point. In Section 5, we carry out a sensitivity analysis of the model parameters. Furthermore, Section 6 presents our discussions alongside the simulation of numerical results. Finally, Section 7 concludes with insights derived from the model and its findings.

## 2. Formulation of Model

We develop a mathematical model to examine the transmission dynamics and prevalence of Tuberculosis, considering five separate epidemiological groups. Within this, individuals who have recovered from the disease are susceptible to re-infection at any time due to interactions with those currently infected with Tuberculosis. The overall population, which interacts uniformly at time *t*, is denoted as *N(t)*. This extensive host population is classified into five epidemiological categories, including susceptible individuals*S(t)*, Individuals who are infected but have not yet reached the stage of being able to transmit the infection (latent/exposed) *E(t)*, as well as those experiencing acute infections*I(t)*, treatment class*T(t)* of tuberculosis carriers and persons who have regained their health *R(t)*. However, these recovered individuals remain at risk of re-infection through contact with tuberculosis-infected individuals.

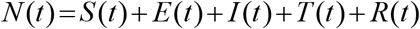

The force of infection is generated by the ratio of the effective contract rate of susceptible individuals with exposed class (*E*)and infected class(*I*), at arate λ given by

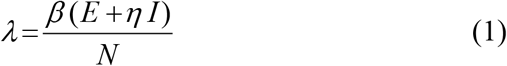

where, *β*denotes the effective contact rate, which refers to the likelihood of contact that can result in tuberculosis infection. Meanwhile, *η* serves as the modification parameter, indicating the infectiousness of acute infection in comparison to the exposed class. In this context, we assume that*η* > 1.

We now describe how we construct the model equations, firstly we construct the susceptible Class 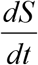, whereindividuals consistof the total constant requirement rate *П*, and this class is decreased by the combination of the force of infection, *λ* and natural death rate, *μ* then the rate at which susceptible individuals change is expressed by

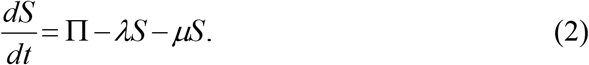

Secondly, we construct a new class where the individuals are not infected but the individuals has some symptoms of infection known as the latent class or exposed class 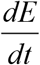. The exposed individuals without any tuberculosis symptoms are generated by the force of infection at the rate *λ* which comes from susceptible individuals. The exposed class is decreased by the progression rate of the exposed class, *κ* and also by the natural death rate, *μ* . Individuals who have been exposed are assumed to initially show no disease symptoms. Thus

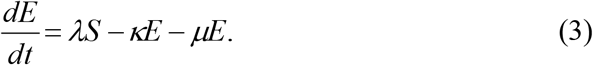

Thirdly, we discuss the individuals who are infected by tuberculosis as acute infection. This population is increased by the progression rate of the exposed class, *κ* and also by a portion of re-infected recovered individuals *fη*_*r*_ *λR*, and decreased by recovery rate, *γ* also treatment rate, *τ*, natural death rate, *μ* and also decreases induced death rate, *δ*_1_. Thus the governing equation is

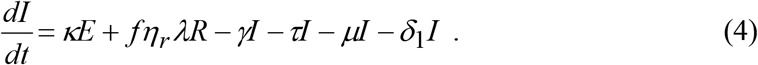

We define the treatment class, 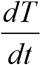 which increases by the treatment rate *τ* of acute infected individuals. This class decreases by the recovery rate, *γ*,natural death rate, *μ* and disease-induced death rate, *δ*_2_ . Therefore, the equation for the treatment individuals becomes:

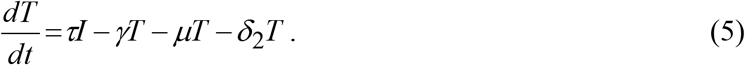

Finally, the recovery individuals increased by the recovery rate, *γ* of acutely infected and treated individuals and decreased by the reinfection of recovered individuals at a rate, *fη*_*r*_ *λR* and natural death rate, *μ* . Hence, the recovery class becomes,

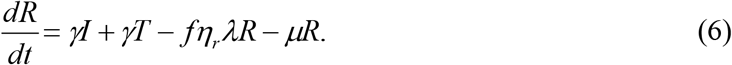

The non-linear differential equations, reflecting the dynamics of tuberculosis transmission, are outlined below, with the relevant variables and parameters specified in Table 1.

**Table 1:** An overview of the model’s variable components.

| Parameters | Description | Values |
| --- | --- | --- |
| $\mu$ | Natural death rate | 0.0121[13] |
| $\lambda$ | Force of infection | - |
| $\eta$ | Acute infection rate | 1.2(Estimated) |
| $\delta_1$ | Diseases induced death rate of acute infected class | 0.1(Estimated) |
| $\delta_2$ | Diseases induced death rate of treatment class | 0.1(Estimated) |
| $f$ | Fraction of re infected recovered individuals | 0.3(Estimated) |
| $\eta_r$ | Re infection rate for R class | 0.05[13] |
| $\kappa$ | Progression rate of exposed class | 0.85(Estimated) |
| $\tau$ | Treatment rate | 0.8(Estimated) |
| $\gamma$ | Recovery rate | 0.23 [13] |
| $\Pi$ | Constant recruitment rate | 2000(Estimated) |
| $\beta$ | Transmission rate | 0.45 [13] |

**Table 2.** Indices of sensitivity of *R*_0_ to the model parameters of TB model:

| Parameters | Indices of sensitivity without treatment ($\tau = 0$) | Indices of sensitivity with treatment ($\tau = 0.8$) |
| --- | --- | --- |
| $\eta$ | 0.224864 | 0.138632 |
| $\kappa$ | -0.24447 | -0.150719 |
| $\beta$ | 0.795104 | 0.294117 |
| $\mu$ | -0.010482 | -0.00646233 |
| $f$ | 0 | 0 |
| $\eta_r$ | 0.0232 | 0.01214 |
| $\gamma$ | -0.0452086 | -0.0278718 |
| $\tau$ | -- | -0.0969454 |
| $\delta_1$ | -0.0196559 | -0.01211182 |
| $\delta_2$ | -0.8923 | -0.5921 |
| $\Pi$ | 0 | 0 |

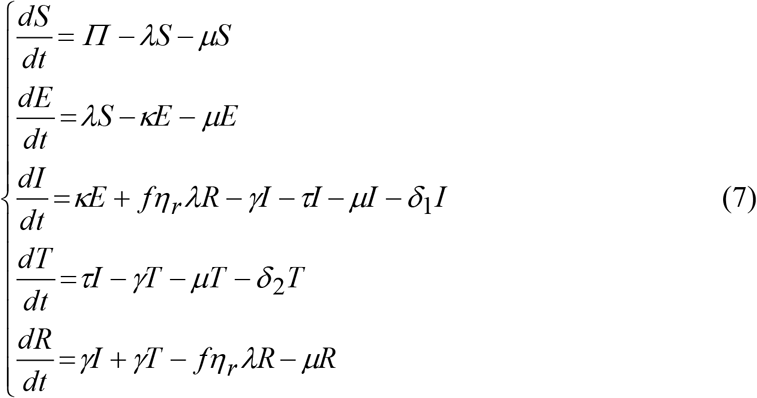

where, *k*_1_ = *κ* + *μ, k*_2_ = *γ* +*τ* + *μ* + *δ*_1_, *k*_3_ = *γ* + *μ* + *δ*_2_, *k*_4_ =□*fη*_*r*_ *λ* + *μ* .

The important features of the model (7) are as follows:

1. Permits the transmission of disease by individuals in the exposed, acute, and removed classes;
2. Individuals who have been exposed display symptoms of the disease at the designated rate and subsequently progress to the acute class;
3. Permits the potential for reinfection in individuals who have previously recovered, at specified rates;
4. Individuals who have been re-infected and subsequently develop symptoms will transition back into the recovered class;

6.Permits treatment (at the specified rate), and temporary release from the disease into acute class.

The model (7) reformed upon the models discussed in previous studies introducing treatment for infected individuals (Beibei Qiu, 2022). Additionally, we incorporatethe standard incidence rate and re-infection in our model. With the modifications made to model (7), we investigate the dynamic characteristics of the model we have proposed.

## 3. Model Analysis

### 3.1 Tuberculosis Free Equilibrium point or DFE

The model (7) exhibits a Disease-Free Equilibrium, which is established by equating the right-hand sides of its equations to zero.

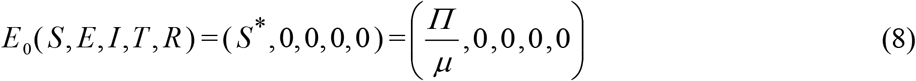

The stability of the DFE, *E*_0_ will be analyzed using the next-generation method (Eka D.A. Ginting, 2024). Presented below are the non-negative matrix *M*, which denotes the new infection terms, and the non-singular M-matrix *V*, which relates to the remaining transfer terms.

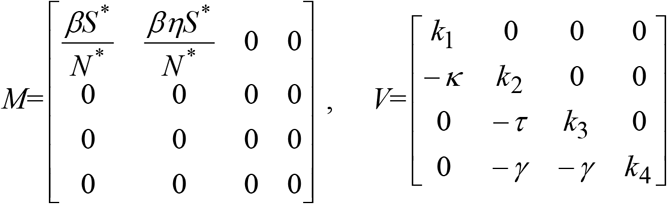

The above matrices at DFE are:

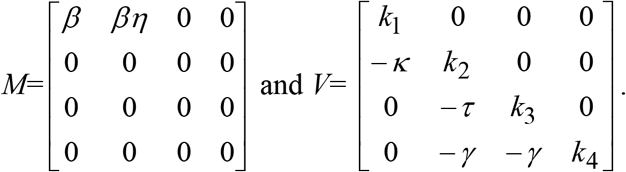

The reproduction number, represented by *R*_0_, is defined as *R*_0_ = *ρ* (*MV* ^−1^), where *ρ* signifies the spectral radius (the largest eigenvalue in absolute value) of the next generation matrix *MV* ^−1^ . Consequently, it can be inferred that

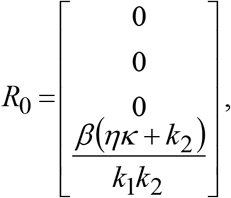

It can also be rewritten as,

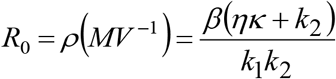

with, *k*_1_ = *κ* + *μ, k*_2_ = *γ* +*τ* + *μ* + *δ*_1_ .

Hence, the result below follows with the help of Theorem 2 (Eka D.A.Ginting, 2024).

#### 3.1.1 LocalStability of Disease Free Equilibrium (DFE)

##### Theorem 1

The model equations (7) arestable locally (asymptotically) if *R*_0_ <1, and unstable if *R*_0_ >1 at DFE, *E*_0_ (Jean Claude Kamgang, 2008).

**Proof:** The demonstration of Theorem 1 relies on the utilization of center manifold theory. (B. Haasdonk, 2021). It is convenient to consider *S* = *x*_1_, *E* = *x*_2_, *I* = *x*_3_, *T* = *x*_4_ and *R* = *x*_5_, so that *N* = *x*_1_ + *x*_2_ + *x*_3_ + *x*_4_ + *x*_5_ .

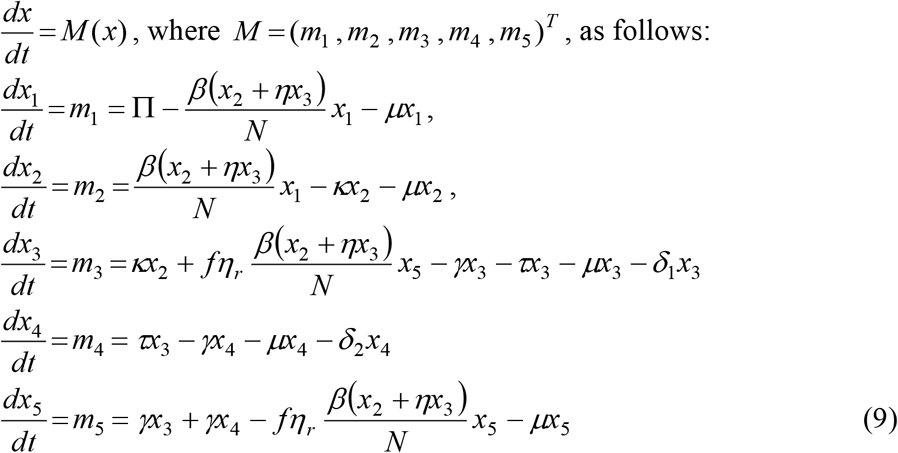

where, *k*_1_ = *κ* + *μ, k*_2_ = *γ* +*τ* + *μ* + *δ*_1_, *k*_3_ = *γ* + *μ* + *δ*_2_, *k*_4_ =□*fη*_*r*_ *λ* + *μ* .

The Jacobian of the system represented by equation (9) or, alternatively, (7),at the DFE, *E*_0_ is expressed as follows

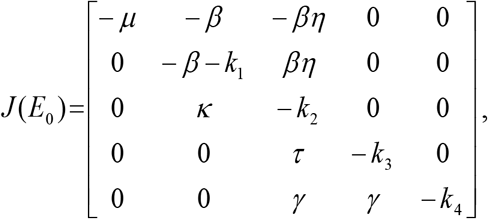

The characteristics equation of the Jacobian matrix is

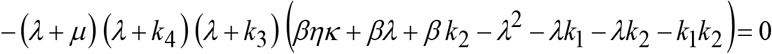

After factorization the characteristic equation becomes,

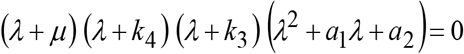

Solving for eigenvalues of *J* (*E*_0_) then we find *λ*_1_ = −*μ, λ*_2_ = −*k*_4_, *λ*_3_ = −*k*_3_ and *λ*^2^ + *a*_1_*λ* + *a*_2_ = 0 from which we get another two eigenvalues, depends on the sign of constant co-efficient of the polynomial.Here *a*_0_ = 1, *a*_1_ = *k*_1_ + *k*_2_ + *β, a*_2_ = *k*_1_*k*_2_ − *β k*_2_ − *βηκ* .

Now taking,

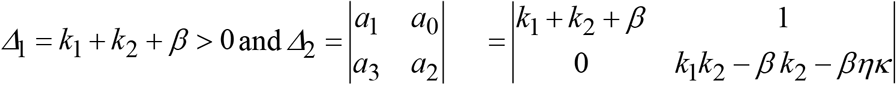

from which it can be shown, (*k*_1_*k*_2_ − *β k*_2_ − *βηκ*) (*k*_1_ + *k*_2_ + *β*) > 0, since (*k*_1_ + *k*_2_ + *β*) > 0, we write, *k*_1_*k*_2_ − *β k*_2_ − *βηκ* > 0 Here first three eigenvalues are negative. According to the Routh-HurtwizCriterion(Jun Zhang, 2024), we get another two eigenvalues are negative if *a*_1_ > 0 and *a*_1_. *a*_2_ > 0, therefore we obtained *a*_1_. *a*_2_ > 0 fulfilled with the conditions,

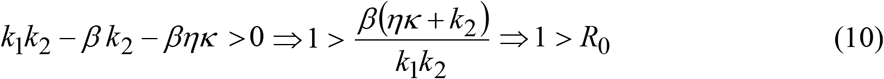

from which it can be shown (as before) that obtained 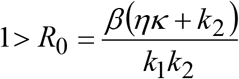, then by using the Routh-Hurtwiz stability condition the characteristic equation has another two negative real roots, it is ensure us that the disease free equilibrium (DFE), *E*_0_ =(*S, E, I*,*T, R*) = 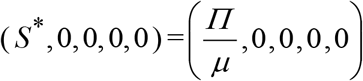 is locally asymptotic stable(Jean Claude Kamgang, 2008).

The importance of Theorem 1 in epidemiology lies in its demonstration that tuberculosis spread can be effectively controlled within a community (when *R*_0_ <1), provided that the initial population sizes in the model remain within the disease-free equilibrium *E*_0_ .

#### 3.1.2 Globalstability of Disease Free Equilibrium (DFE)

To demonstrate the global stability of the DFE in model (7), we will apply the Castillo-Chavez theorem (Yujiang Liu, 2016). To proof we rewrite the model equation as:

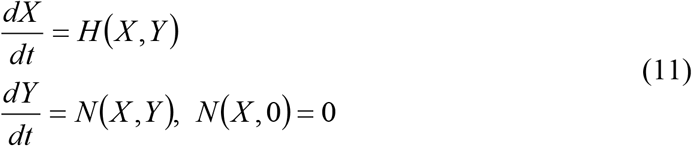

where, *X* = *S* and *Y* = (*E, I*,*T, R*), with the components of 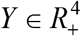 denoting the infected population. The disease-free equilibrium is now denoted as,

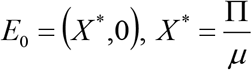

The condition must be met a local asymptotic stability 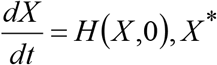 is the DFE which is globally asymptotically stable(Junli Liu, Global stability for a tuberculosis model, 2011).

We consider, 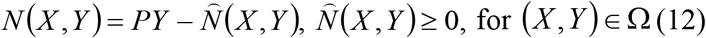, for (*X, Y*)∈ Ω (12) where, *P* = *D*_*z*_ *N* (*X* ^*^, 0) is a off-diagonal non-negative M matrix and *Ω* makes biological sense for the model where the model is exist. If the system satisfies the above conditions then the following theorem is hold.

##### Theorem 2

If a fixed point *E*_0_ = (*X* ^*^,0)of any state is present, it will be GAS as an equilibrium of the system of equationswhen satisfied. *R*_0_ < 1, provided that the conditions in (12) are

**Proof:** Equation (11),(12)implies

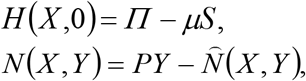

where,

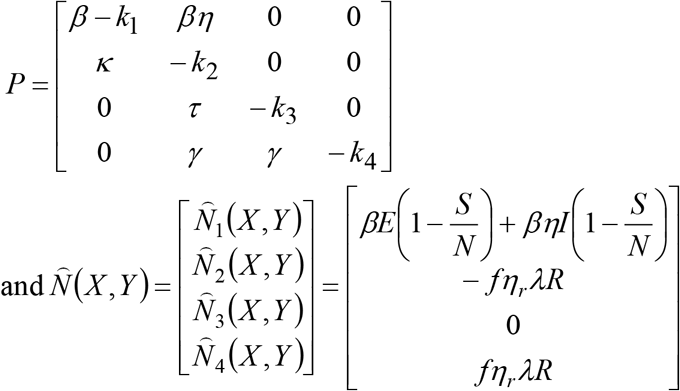

From the above matrix we see that 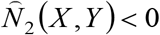 which can not satisfies the condition (12). So, 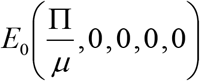 the disease-free equilibrium may or may not exhibit global asymptotic stability under certain conditions *R*_0_ < 1 (Junli Liu, Global stability for a tuberculosis model, 2011).

## 4. Uniqueness and Presence of Endemic Equilibrium Point (EEP)

This section assesses the potential presence and stability of endemic (positive) equilibria within model (7). It will focus on scenarios where at least one of the infected components is greater than zero, assuming that there is no external re-infection (Youngjoon Cha, 2022).

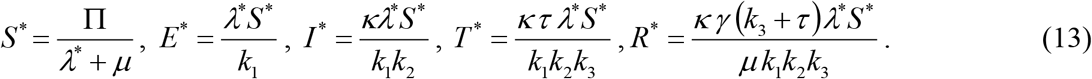

Positive endemic equilibrium points exist solely when *R*_0_ >1 represented by the notation, *λ*^*^ is applicable at these points.

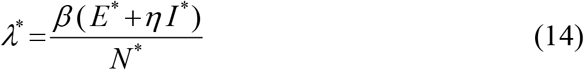

Substituting the endemic equilibrium point in the expression (14) we have,

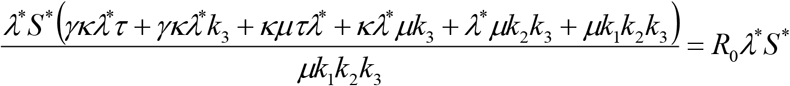

Dividing both sides by *λ*^*^*S* ^*^ (it is mentioned at *λ*^*^*S* ^*^ ≠ 0) becomes

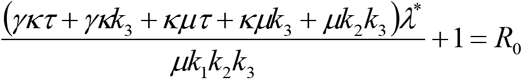

After some simplification we get

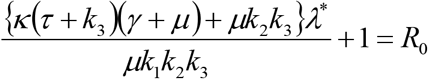

Which implies, *Pλ*^*^ +1 = *R*_0_, where 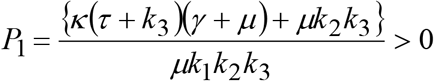 Therefore, 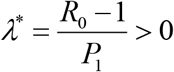, whenever *R*_0_ > 1 .

Thus the following lemma is investigated(Oyovwevotu, 2021).

### Lemma 1

*The model (7) has a unique endemic equilibrium point(EEP) with η*_*r*_ = 0 *whenever R*_0_ > 1 .

### 4.1 Local Stability analysis of the EEP

#### Theorem 3

*If R*_0_ > 0 *then the EE point is locally asymptotic stable*.

**Proof:** To illustrate the local stability analysis of the EEP we will use the theory of Center Manifold (Tao Zhao, 2020) with no exogenous re-infection (that is *η*_*r*_ = 0) occurs. Firstly we assume the equilibrium point at bifurcation point, when *R*_0_ = 1 and analyze the dynamics with *β* = *β* ^*^ .

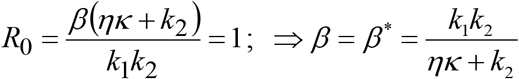

The Jacobian matrix at *β* ^*^ is,

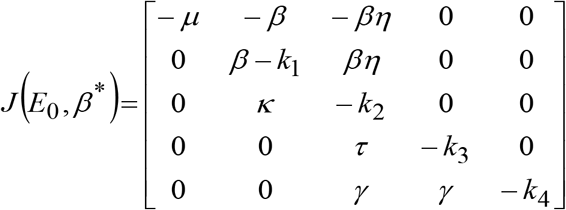

where

*J* (*E*_0_, *β* ^*^).*w* = 0 .The linear equations of the Jacobianmatrix given by

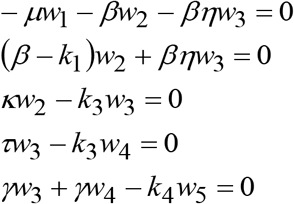

From which we get the right eigenvectorssuch as,

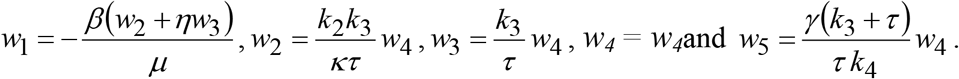

#### Computations of the bifurcation coefficients *a* and *b*

To calculate *a*, partial derivatives in (9) gives :

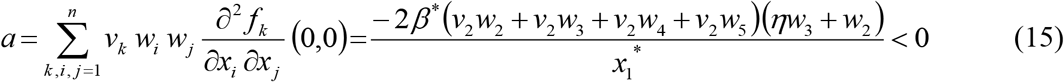

To calculate *b*, it can be shown that

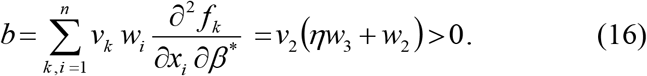

By substituting the corresponding left and right eigenvectors in (15) and (16) confirmed bifurcation coefficients *a* is negative and *b* is positive. So according to the theorem of (Hao Shen, 2023), we can concludeabout uniqe EEP is LAS if *R*_0_ > 1.

##### Theorem 4

*The model demonstrates a unique endemic equilibrium that is locally asymptotically stable under specific conditions R*_0_ > 1 . *Consequently, when particular parameters a* < 0 *and b* > 0 *met, the endemic equilibrium is validated as locally asymptotically stable in accordance with the principles of Center Manifold Theory (Herbert W. Hethcote, 2022)*.

### 4.2 Global Stability of Endemic Equilibrium Point (EEP)

The global asymptotic stability of the EEP (HosseinMohebbi, 2018), *E*_1_ =(*S* ^*^, *E*^*^, *I* ^*^,*T* ^*^, *R*^*^) is examined in relation to the reduced model (9) under the circumstances of re-infection*η*_*r*_ = 0 and *δ*_1_ = 0, *δ*_2_ = 0 . By establishing the appropriate conditions 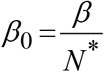, it can be demonstrated that the reproduction number associated with the reduced model (9) is validwith *η*_*r*_ = 0 and *δ*_1_ = 0, *δ*_2_ = 0, denotedby *R*_01_, is given by

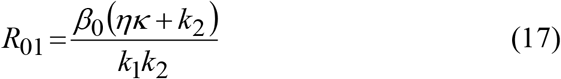

where, *k*_1_, *k*_2_ are defined in section 3 with *η*_*r*_ = 0 and *δ*_1_ = 0, *δ*_2_ = 0 . Furthermore, using the approach in section4, it can be shown that the reduced system (9), has a unique EEP,of the form

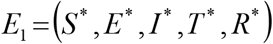

where, *S* ^*^ > 0, *E*^*^ > 0, *I* ^*^ > 0,*T* ^*^ > 0, *R*^*^ > 0 whenever *R*_01_ > 1.

#### Th^m^ 5

*The unique EEP, E*_1_, *of the reduced model (9) with η*_*r*_ = 0 *and δ*_1_ = 0, *δ*_2_ = 0, *is GASwhenever R*_01_ > 1.

**Proof:** Let *η*_*r*_ = 0 and *δ*_1_ = 0, *δ*_2_ = 0 and *R*_01_ > 1, Examine the subsequent derivative of the non-linear Lyapunov function as presented by(Huijuan Li, 2020):

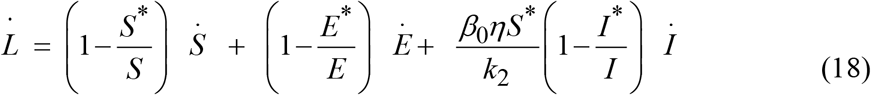

Now,

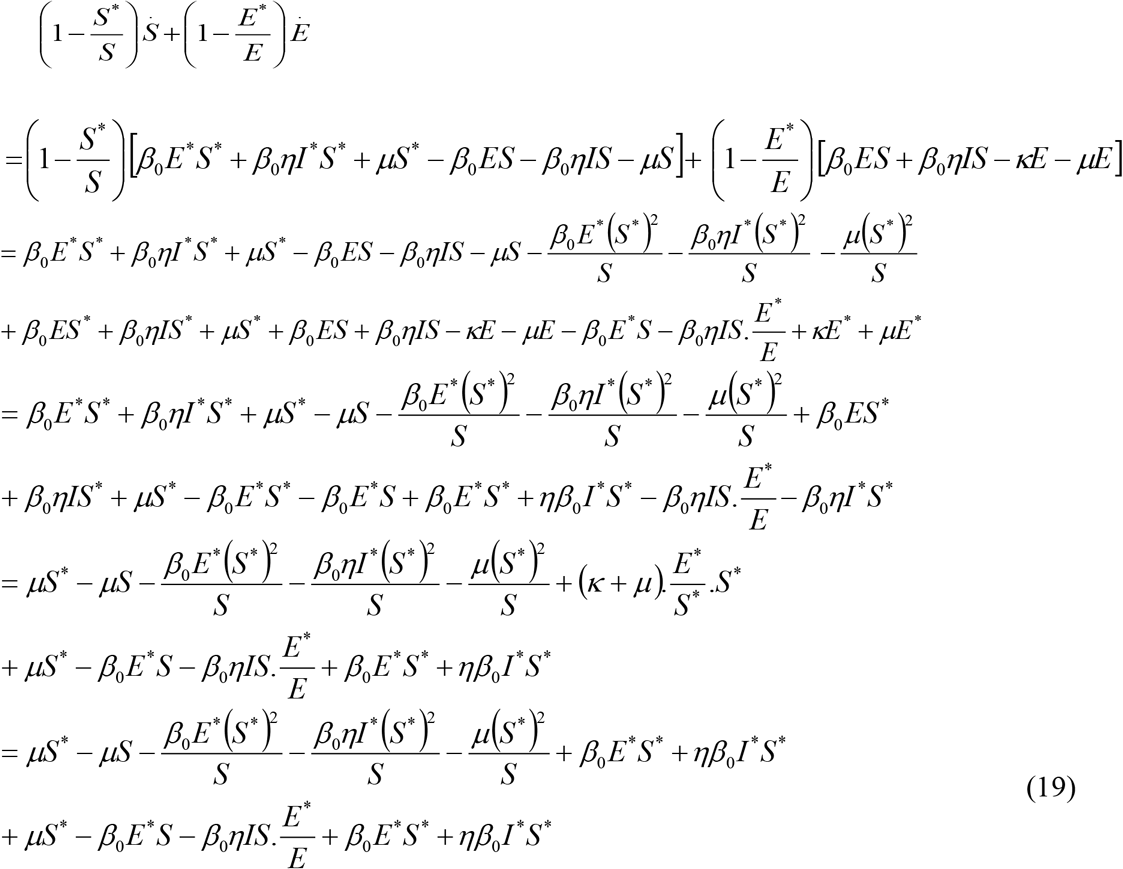

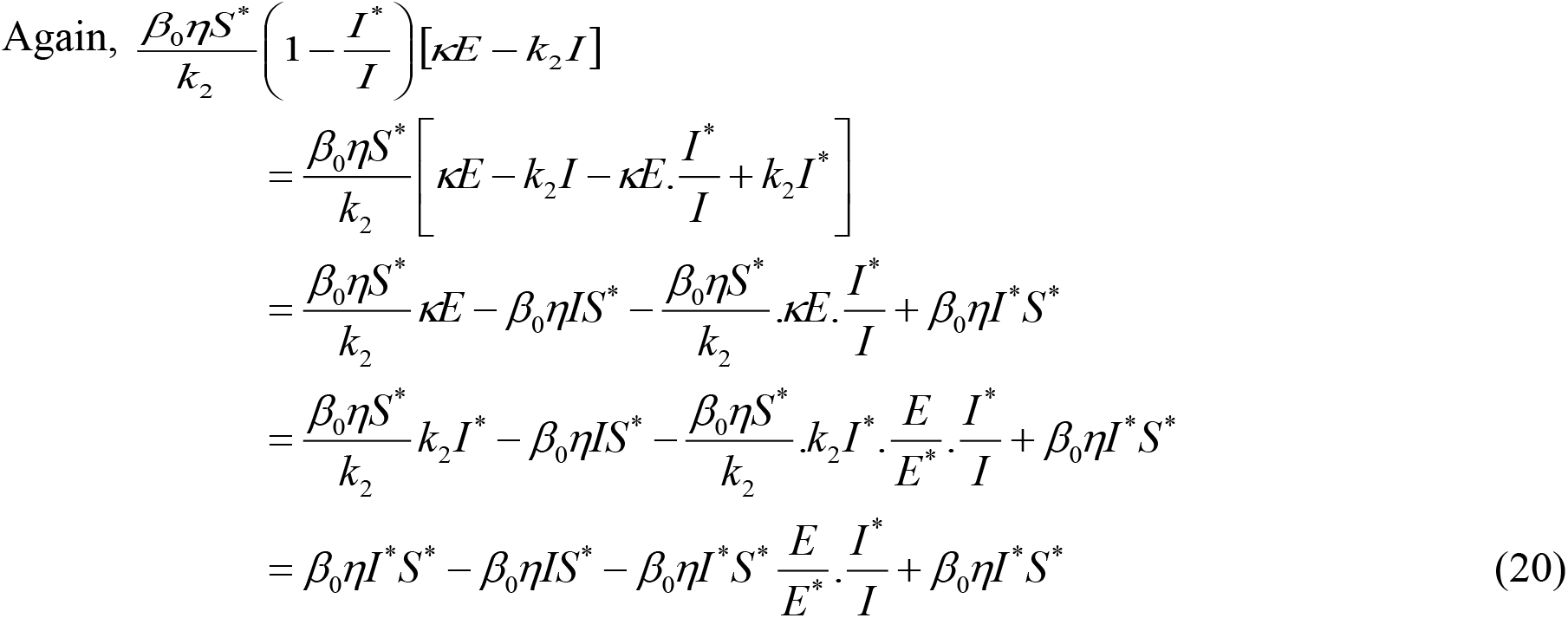

Adding (19) and (20) and substituting in (18) for simplification we have,

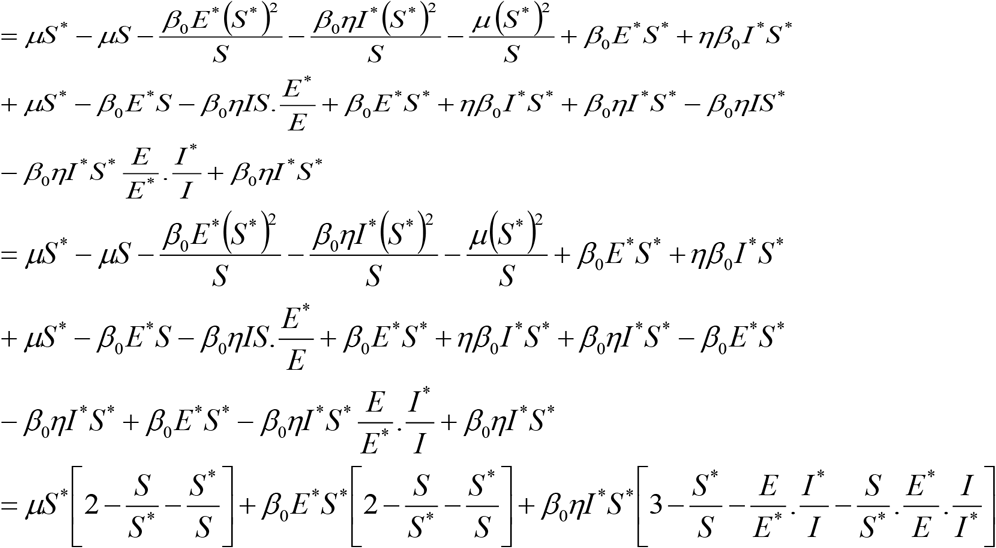

Because the arithmetic mean is greater than the geometric mean, it follows that

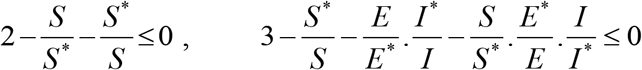

So that Ŀ≤ 0 for

*R*_01_ > 1. Hence, *L* is a Lyapunov function of the system (9), with *η*_*r*_ = 0 and *δ*_1_ =*δ*_2_ = 0 . Consequently, utilizing the Lyapunov function and aditionallywith LaSalle’s Invariance Principle (Jinhuan Wang, 2008), it can be concluded that every solution to the equations in the reduced model (9) converges to *E*_1_ as t → ∞ for *R*_01_ > 1.

## 5. Sensitivity Analysis

Sensitivity analysis serves as a technique to identify the essential parameters that are vital for comprehending the dynamics of a model (Damien A. Fordham, 2016). In the realm of mathematical epidemiology, this method is crucial for identifying parameters that are sensitive and pertinent to disease management. By performing sensitivity analysis on the parameters of the model, we can identify those that have a significant effect on the model’s dynamics. Specifically, we compute sensitivity indices for the basic reproduction number using the parameters specified in the model (7) to ascertain which parameters notably affect the reproduction number and the transmission of the disease. This methodology is grounded in the work of Md. Samsuzzoha (2013). Sensitivity indices offer a way to measure the relative change in a state variable resulting from variations in parameters. The normalized forward sensitivity index of a variable concerning a parameter is defined as the ratio of the relative change in the variable to the relative change in the parameter. A negative sensitivity index indicates an inverse relationship between the parameters and the reproduction number (ZhaoxiaXu, 2024). In such cases, the focus is placed on the absolute value of the sensitivity index.

### 5.1 Sensitivity Indices of *R*_0_

To analyze the sensitivity of *R*_0_ (Xiunan Wang, 2019)we defined an explicit formula for *R*_0_ as,

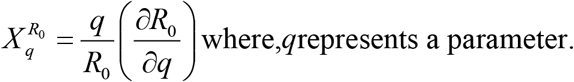

The above table illustrates that all the parameters’ behavior shows a positive or negative impact on the rate of reproduction number, *R*_0_ . From the above table, we observe that*η* and *β* that is the infection rate and the transmission rate are more positively sensitive parameters of the treatment effect(Xiunan Wang, 2019). We can understand that if we increase the value of those two parameters then the value of *R*_0_ is also increased because they are positively sensitive with *R*_0_ .

On the other hand, other parameters such as the progression rate of the exposed class, recovery rate and disease-induced death rate of the infected class (*κ*,*γ*,*δ*_1_) are the most negatively sensitive with *R*_0_ . That is if we increase the value of these three parameters, there is a negative impact on *R*_0_ . But when we decrease the value of these parameters then there is a positive impact on *R*_0_ when treatment is not applied (C. Edouard, 2016).

A sensitivity analysis of *R*_0_ was conducted, showing that parameters such as infection rate, transmission rate, and rate of movement from exposed to active TB, *η* are directly related to *R*_0_ . Furthermore, the study reveals that *R*_0_ decreases with higher recovery rate, *γ□*andtreatment rate,*τ* and also indicated that improved treatment can lead to increased recovery and decrease TB infections, reducing overall mortality rates. Besides the other parameters are less sensitive in terms of disease burden(Susanne F. Awad, 2020).

### 5.2 The coefficient of partial rank correlation (PRCC)

The Latin Hypercube Sampling and Partial Rank Correlation Coefficient (PRCC) sensitivity analysis represent a highly effective approach frequently employed in uncertainty analysis to examine all parameters of a model (J.C. Helton, 2003). The application of partial rank correlation is prevalent in sensitivity studies. This method is noted for its superior capability in identifying the sensitivity of parameters that exhibit strong monotonicity yet are relatively nonlinear (Abdul Malek, 2024). PRCC serves as a method for sensitivity assessment, computing the partial rank correlation coefficient between the inputs and outputs of the model (Sorokin,2023).

For the Tuberculosis model (7), the sensitivity index shows that the acute infection rate (*η*), disease-induced death rates of acute and treated classes (*δ*_1_ and *δ*_2_), treatment rate (*τ*), recovery rate (*γ*) and transmission coefficient rate (*β*) are the most influential parameters, with corresponding indices of 0.5555, − 0.5560, − 0.8472, − 0.4168, − 0.9744 and 0.9579 respectively, as indicated in Figure 4. Overall, the result summarizes that if the transmission rate (*β*) increases unboundedly, it will be difficult to maintain the disease outbreak. On the other hand, the treatment rate (*τ*)□and the recovery rate (*γ*)□may be the most effective influential parameter to reduce the diffusion of disease transmission from the community.

**Figure 1.**
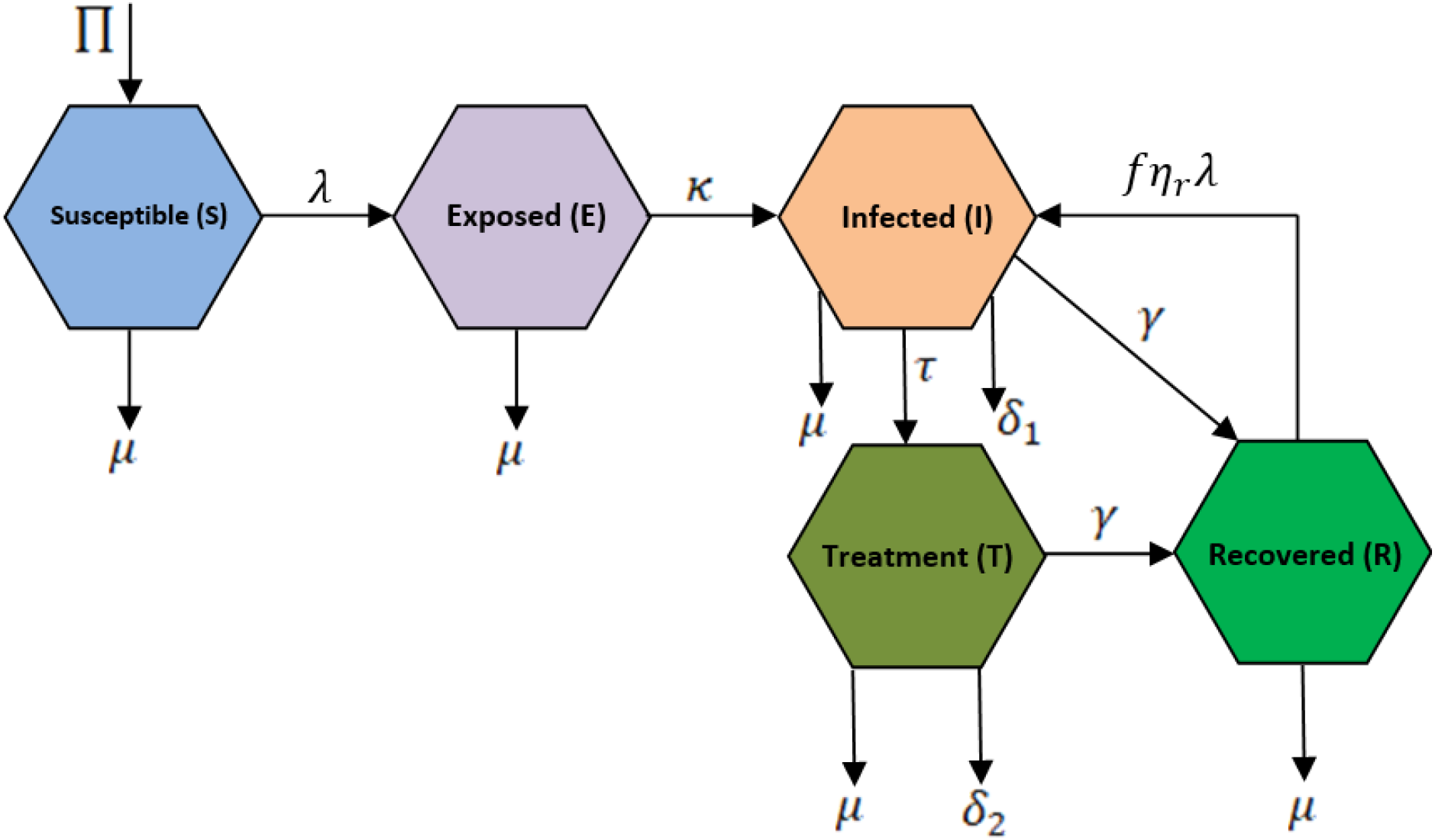
Model diagram of TB transmission

**Figure 2.**
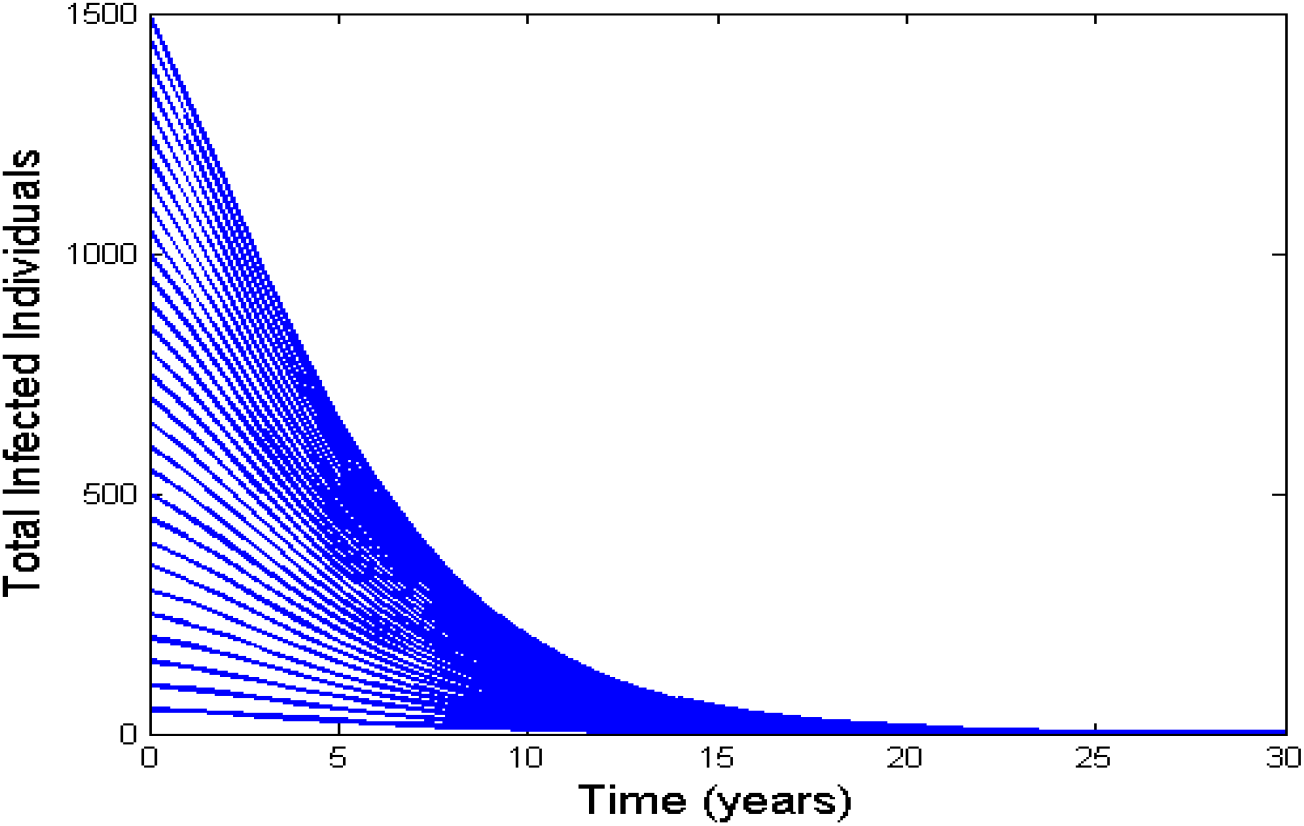
This Time series plot illustrates that the DFE is locally asymptotically stable (LAS) when *R*_0_ = 0.8304 < 1 .

**Figure 3.**
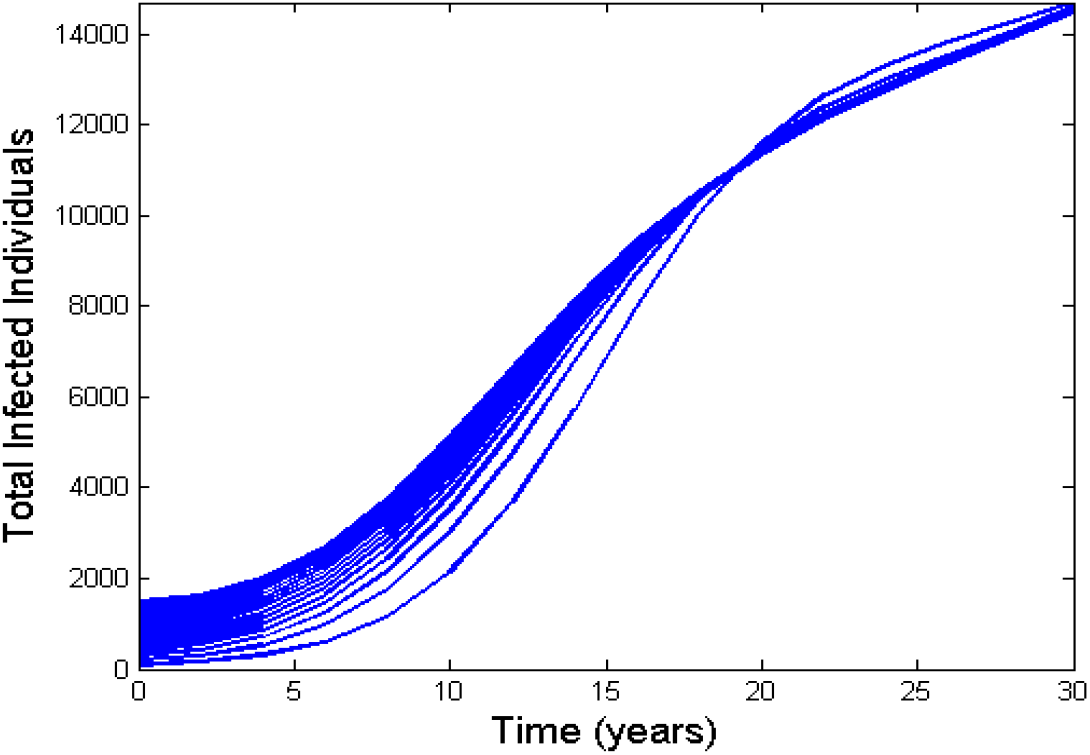
This Time series plot illustrates that the unique EEP is locally asymptotically stable (LAS) whenever *R*_0_ =1.5593 > 1.

**Figure 4.**
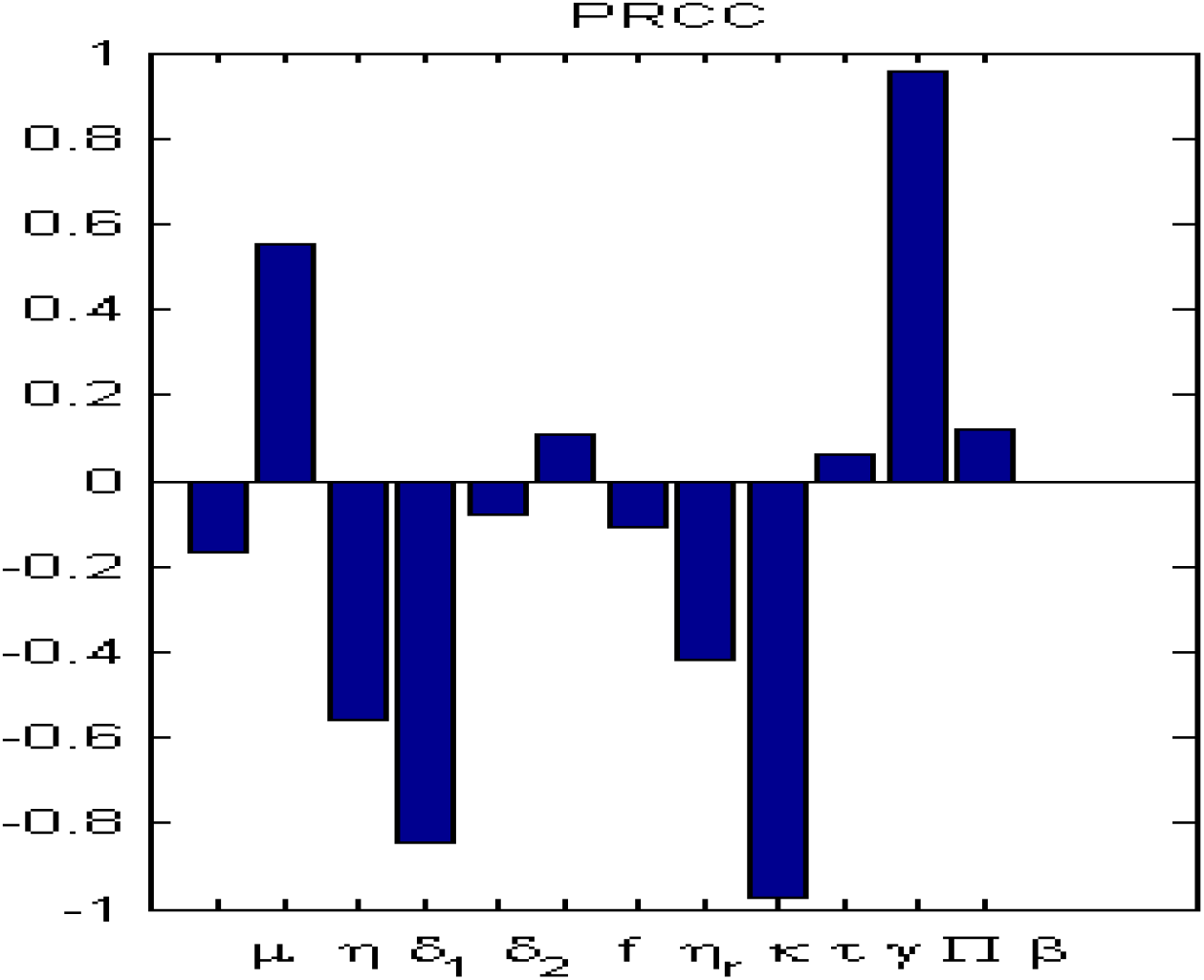
PRCC index of the parameters of the TB model.

## 6. Computational Simulations and Analysis

This section presents several theoretical findings are supported by the computational simulations of the model (7) using different parameters given in Table 1, parameter values computations are either (Zukowski, 2007) or arbitrarily chosen to estimate the desired results.

Figure 5 indicates treatment has a positive impact since it increases at a slow rate in comparison to the other classes when *R*_0_ > 1. Also, the infection rate increases very rapidly when the treatment is not applied.

**Figure 5.**
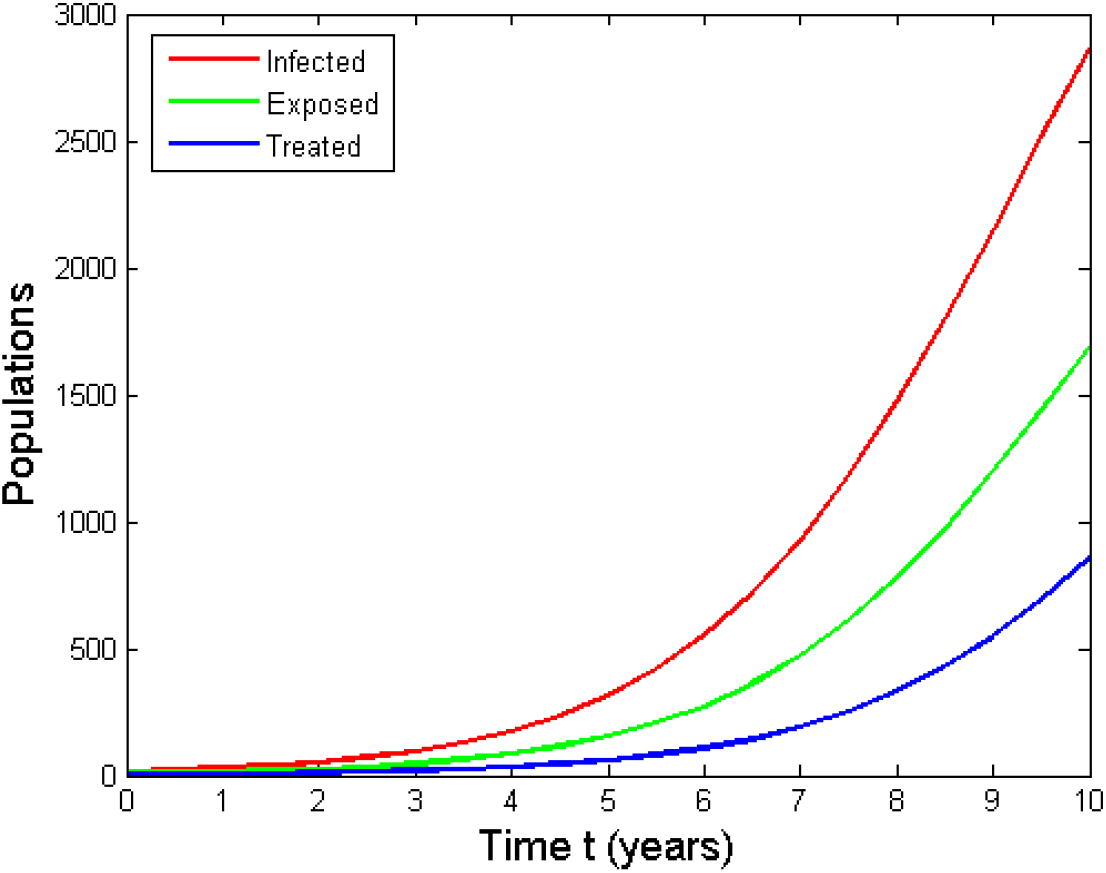
Dynamic transmission of infected classes whereas *R*_0_ > 1 .

In Figure 6(a), the solution curves of different classes of population approach a locally stable DFE when the set of parameter values is (*β* = 0.25,*κ* = 0.85,*γ* = 0.23, *μ* = 0.014)whereas *R*_0_ = 0.4613 < 1 . This occurs only when the contact rate is low and while the recovery rate is expanded thus the reproduction number is not more than one.

**Figure 6(a):**
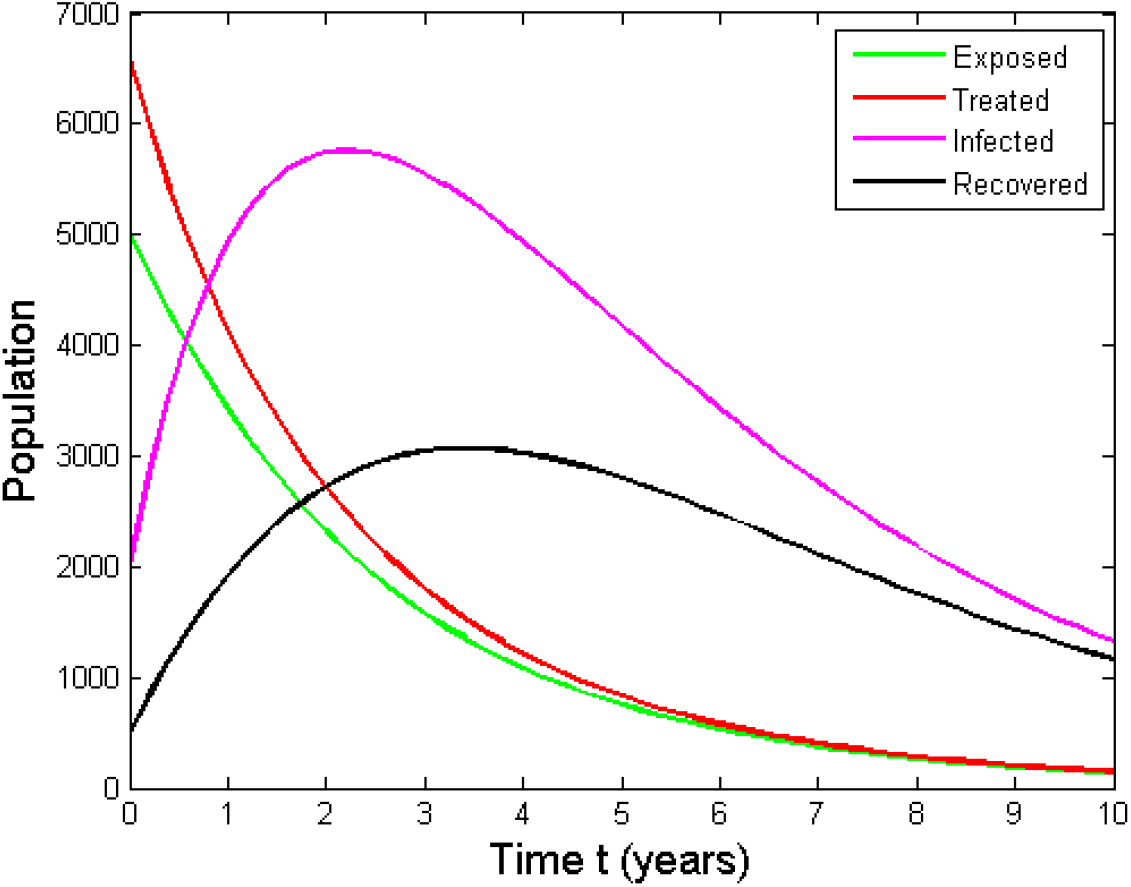
Solution curves of TB model with parameters *β* = 0.25,*κ* = 0.85,*γ* = 0.23, *μ* = 0.014 when *R*_0_ < 1 .

**Figure 6(b):**
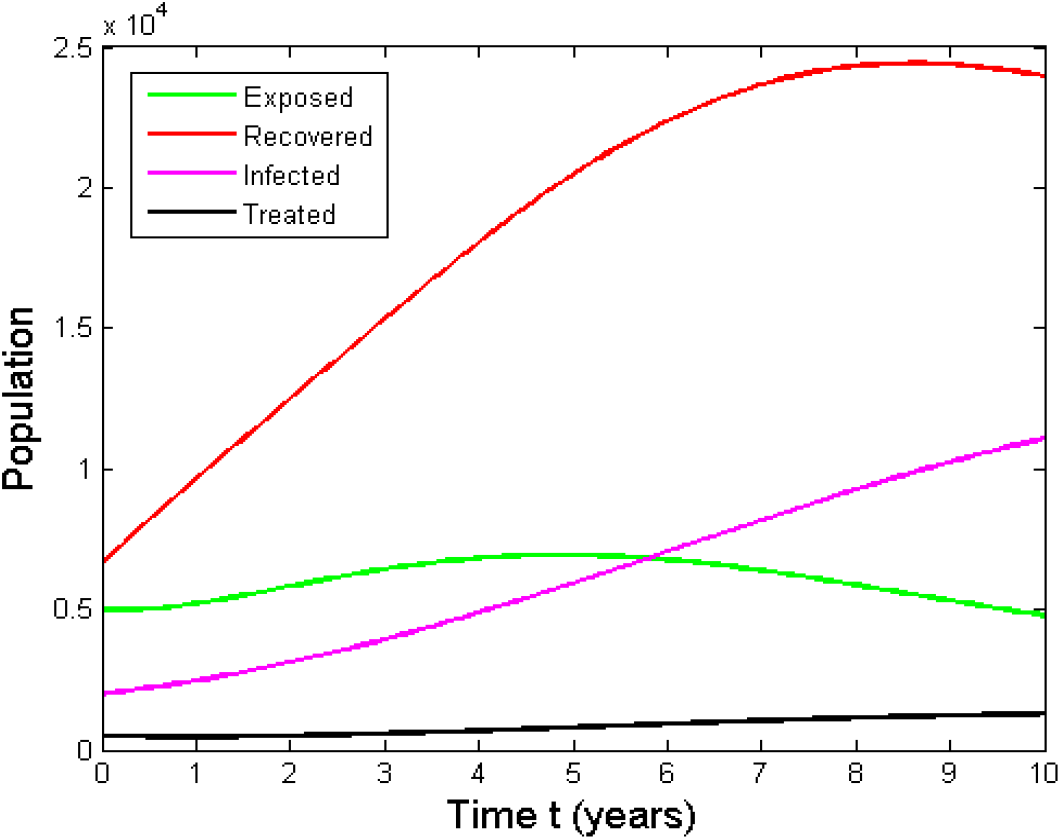
Solution curves of TB model with parameters *β* = 0.65,*κ* = 0.95,*γ* = 0.02, *μ* = 0.005 when *R*_0_ > 1.

In Figure 6(b), the solution curves of different classes of population approach a locally stable EEP when the set of parameter values is (*β* = 0.65,*κ* = 0.95,*γ* = 0.02, *μ* = 0.005) whereas *R*_0_ = 3.08 > 1 . Numerical computation of the model (7) showing with a high contact rate and less recovery rate, when the reproduction number is more than one.

### 6.1 Effect of Transmission co-efficient (*β*)

From Figure 7, it is evident that lower values of *β* indicate a high effectiveness of disease control measures in managing contacts with infected individuals or contaminated sources. Conversely, higher values of *β* signify a low effectiveness of disease control. For the numerical computation of model (7), three arbitrary effectiveness levels of TB control measures concerning contact rates are taken as:

1. Low effectiveness level of disease burden control where the contact rate measures at *β* = 0.4,
2. Moderate effectiveness level of disease burden control where the contact rate measures at *β* = 0.06,
3. High moderate effectiveness level of disease burden control where the contact rate measures at *β* = 0.002 .

**Figure 7.**
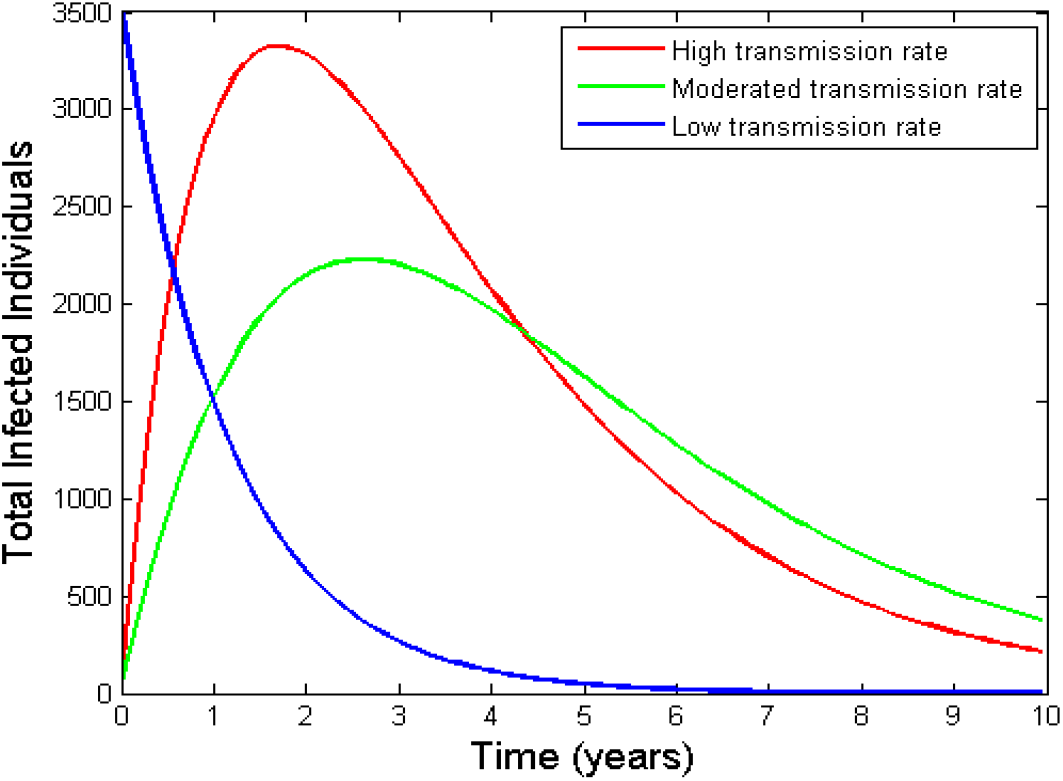
Simulation of the model (7) using different transmission rate *β* = 0.4, *β* = 0.06, *β* = 0.002 for effectiveness levels of TB control when *R*_0_ < 1.

### 6.2 Effect of treatment rate (*τ*) on TB control

Numerical computations illustrated in Figure 8 evaluate the influence of treatment strategies with varying effectiveness levels in tuberculosis (TB) control. The analysis considers three distinct effectiveness levels of TB control measures corresponding to different treatment intensities, such as:

1. The low efficacy of the treatment (*τ* = 0.008) is associated with a swift rise in infections with *β* = 0.4 .
2. The moderated efficacy of the treatment (*τ* = 0.5) resulting in a gradual reduction in the infection with *β* = 0.06 .
3. The high efficacy of the treatment (*τ* = 0.8) which is associated with a gradual rise in infection rates with *β* = 0.002 .

**Figure 8.**
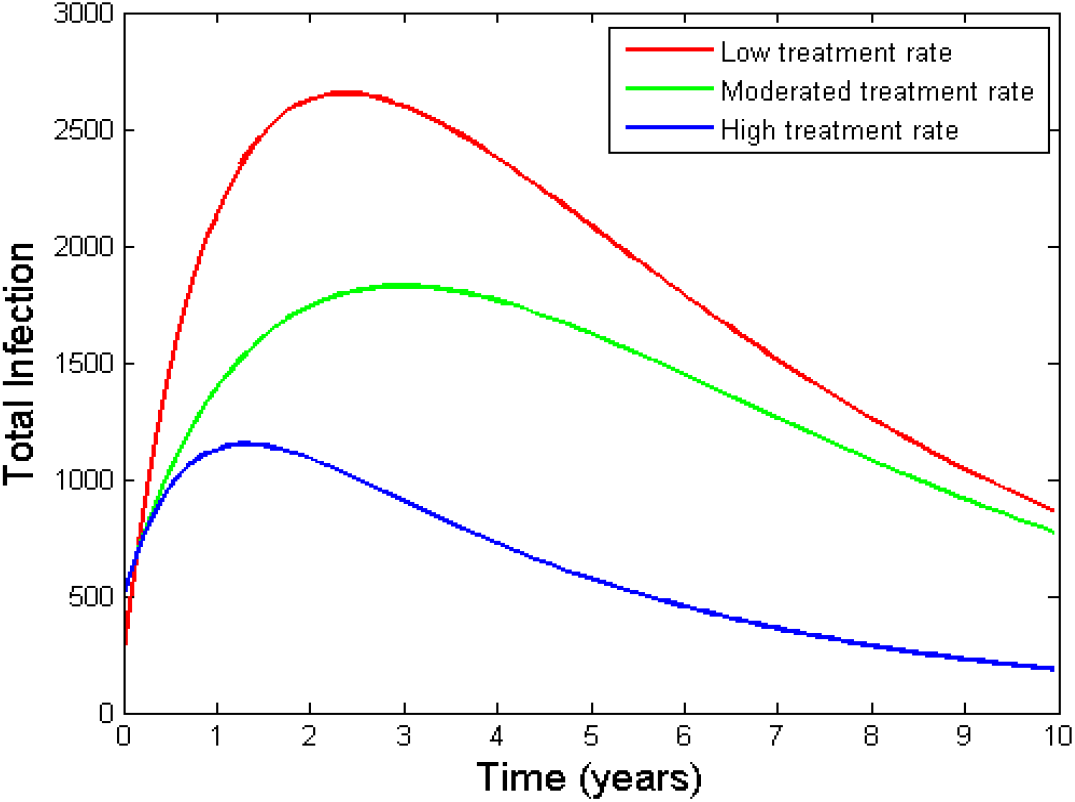
Simulation of the model (7) using different treatment rate *τ* = 0.8,*τ* = 0.5,*τ* = 0.08 to show transmission effect when *R*_0_ < 1.

## 7. Conclusions

A deterministic treatment-structured model was developed to examine tuberculosis transmission, persistence, and control. The population was divided into susceptible, exposed, actively infected, treated, and recovered classes, and reinfection among recovered individuals was incorporated. Through the inclusion of a distinct treatment compartment, the epidemiological effects of treatment initiation, recovery during treatment, treatment-associated mortality, and reduction of infectious burden were represented more explicitly than would be possible when treatment is included only as a transition parameter. The model was shown to possess biologically meaningful nonnegative solutions within a feasible region. A disease-free equilibrium and an endemic equilibrium were derived, and the basic reproduction number was established as the principal threshold governing tuberculosis invasion and persistence. Local asymptotic stability of the disease-free equilibrium was obtained when . Thus, transmission was predicted to decline when the reproduction threshold was maintained below unity. When, sustained transmission and the existence of a unique endemic equilibrium were indicated. Under the reduced-model assumptions adopted in the analysis, local and global stability properties of the endemic state were also established. The sensitivity findings showed that infection- and transmission-related parameters produced the strongest positive effects on the reproduction number and disease burden. Conversely, treatment, recovery, and relevant removal parameters were found to exert protective effects. The PRCC analysis further identified the transmission coefficient, progression to active infection, treatment rate, recovery rate, and disease-induced mortality parameters as major determinants of model behavior. The numerical results indicated that increased transmission intensified the epidemic, whereas greater treatment effectiveness substantially reduced the infectious population. It was therefore demonstrated that treatment should be regarded not only as an individual clinical intervention but also as a population-level control mechanism. Tuberculosis elimination was shown to require a combined strategy in which effective treatment, rapid movement of infectious patients into care, improved recovery, reduced transmission, and prevention of reinfection are jointly strengthened. The proposed framework may therefore be used to support treatment-oriented policy evaluation and future intervention modeling.

Overall, the suggested model is a complete tool to explain the transmission of tuberculosis, evaluate treatment-oriented control methods and assist evidence-based decision making in public health. The model includes latent infection, active disease development, treatment dynamics, and reinfection processes and gives useful information for creating successful methods to reduce TB prevalence and restrict long-term disease persistence.

## Data Availability

All the relevant data are disclosed in the article.

## CRediT authorship contribution statement

**Jannatun Nayeem:** Writing – review & editing, Supervision, Software, Methodology, Data curation. **Md. Abu Salek**: Writing – original draft, Project administration, Methodology, Funding acquisition, Formal analysis, Data curation, Conceptualization. **M. H. A. Biswas:** Supervision, Resources, Methodology. **M. Humayun Kabir:** Writing – review & editing, Methodology, Formal analysis, Conceptualization.

